# Anonymizing medical documents with local, privacy preserving large language models: The LLM-Anonymizer

**DOI:** 10.1101/2024.06.11.24308355

**Authors:** Isabella C. Wiest, Marie-Elisabeth Leßmann, Fabian Wolf, Dyke Ferber, Marko Van Treeck, Jiefu Zhu, Matthias P. Ebert, Christoph Benedikt Westphalen, Martin Wermke, Jakob Nikolas Kather

**Affiliations:** Else Kroener Fresenius Center for Digital Health, Technical University Dresden, Dresden, Germany; Department of Medicine II, Medical Faculty Mannheim, Heidelberg University, Mannheim, Germany; Department of Medical Oncology, National Center for Tumor Diseases (NCT), Heidelberg University Hospital, Heidelberg, Germany; DKFZ Hector Cancer Institute at the University Medical Center, Mannheim, Germany; Molecular Medicine Partnership Unit, European Molecular Biology Laboratory, Heidelberg, Germany; Department of Medicine I, University Hospital Dresden, Dresden, Germany; Department of Medicine III, University Hospital, LMU Munich, Marchioninistraße 15, 81377, Munich, Germany; German Cancer Consortium (DKTK), Partner Site Munich, Munich, Germany; Comprehensive Cancer Center (CCC Munich LMU), LMU University Hospital Munich, Munich, Germany

## Abstract

**Background:** Medical research with real-world clinical data can be challenging due to privacy requirements. Ideally, patient data are handled in a fully pseudonymised or anonymised way. However, this can make it difficult for medical researchers to access and analyze large datasets or to exchange data between hospitals. De-identifying medical free text is particularly difficult due to the diverse documentation styles and the unstructured nature of the data. However, recent advancements in natural language processing (NLP), driven by the development of large language models (LLMs), have revolutionized the ability to extract information from unstructured text.

**Methods:** We hypothesize that LLMs are highly effective tools for extracting patient-related information, which can subsequently be used to de-identify medical reports. To test this hypothesis, we conduct a benchmark study using eight locally deployable LLMs (Llama-3 8B, Llama-3 70B, Llama-2 7B, Llama-2 70B, Llama-2 7B “Sauerkraut”, Llama-2 70B “Sauerkraut”, Mistral 7B, and Phi-3-mini) to extract patient-related information from a dataset of 100 real-world clinical letters. We then remove the identified information using our newly developed LLM-Anonymizer pipeline.

**Results:** Our results demonstrate that the LLM-Anonymizer, when used with Llama-3 70B, achieved a success rate of 98.05% in removing text characters carrying personal identifying information. When evaluating the performance in relation to the number of characters manually identified as containing personal information and identifiable characteristics, our system missed only 1.95% of personal identifying information and erroneously redacted only 0.85% of the characters.

**Conclusion:** We provide our full LLM-based Anonymizer pipeline under an open source license with a user-friendly web interface that operates on local hardware and requires no programming skills. This powerful tool has the potential to significantly facilitate medical research by enabling the secure and efficient de-identification of clinical free text data on premise, thereby addressing key challenges in medical data sharing.

## Background

The digitization of medical records, which include clinical letters, reports and various other forms of patient data^1^, has substantially grown in the past years.^2,3^ The Electronic Health Record (EHRs) is moving towards unified digital repositories that encapsulate each patient’s complete healthcare journey, with the goal of creating integrated platforms accessible to healthcare professionals on a national scale, and even between international healthcare systems, for example within the European Union.^4^ This centralization of data potentially enhances clinical utility and facilitates more seamless and continuous healthcare for providers and patients. This transition opens new avenues for a text data-driven approach to medical research^5^ by enabling the systematic extraction of structured information from the growing volume of digital text-based hospital data using natural language processing (NLP) tools.

Recently, the introduction of Large Language Models (LLMs) has markedly facilitated and extended the capabilities of NLP in medicine.^6^ While developing conventional NLP software requires specific training on annotated text in order to retrieve information, generalist LLMs can solve similar tasks without further fine-tuning. Contemporaneous LLMs possess some medical knowledge^7^ and demonstrate an excellent performance for extracting structured information from unstructured medical texts.^8–11^ Simultaneously, the relevance of applying AI algorithms to analyze diverse types of unstructured medical data, such as imaging, text, and genomic data, is increasing. This trend is culminating in the development of general medical foundation models.^12^ All of these new technologies rely on accessible, real-world, large-scale medical datasets.^12,13^

The integration and use of such large-scale datasets present multiple challenges, among them the most essential: ensuring patient privacy. The sensitive nature of patient data precludes the indiscriminate sharing of free text between institutions, sometimes even within one institution, and carries the risk that potential breaches could compromise personally identifiable information (PII) during research procedures. Anonymizing patient data before using it for research is therefore the gold standard to preserve privacy in research, but automated anonymization of medical documents as clinical letters is not trivial.^14,15^ Medical documents exist in a plethora of formats that vary between hospitals and even departments, making it challenging to find a universal solution for anonymization. Current approaches often rely on time-consuming, expensive and imprecise manual work^16^ or Named Entity Recognition (NER) keyword search requiring costly software.^17^

We developed a new anonymization tool based on locally deployable LLMs that facilitates the de-identification of patient data at the origin of its digital storage; on local computer hardware of hospitals and healthcare providers. Our browser-based frontend provides a pipeline incorporating an LLM to process text-based medical records, including multiple file formats such as Portable Document Format (PDF), Text File Documents (TXT) and Microsoft Word documents (Word) containing patient reports. The tool extracts the text data from these files, iteratively analyzes it using the LLM to identify PII and removes it in a second step. The output contains deidentified reports along with a table cataloging the extracted PII.

We systematically and quantitatively evaluated the capabilities of several LLMs which can be employed at the point of care, including Llama-3 and Llama-2-based models as well as Mistral models. Our solution represents a practical advance in privacy-preserving technologies for healthcare data management - a step towards using the vast potential of digital health records for research and clinical improvement without compromising the nature of patient privacy. Crucially, this tool is designed for simple accessibility, requiring no advanced programming skills from its users and it is released as an open source tool.

## Methods

### Aims and Research Question

The goal of our study was to investigate the feasibility of using an LLM-based pipeline for anonymizing medical documents in a zero-shot way, without the need for any training of a dedicated LLM. We benchmark this pipeline on real medical text data and various locally deployable LLMs. In addition, we developed a user-friendly, open-source frontend that allows anonymization of medical documents, the evaluation of the anonymization with an annotated dataset, and easy document review.

### Benchmarking an LLM-based pipeline for anonymization

#### Data collection

The data used for the benchmarking experiments included n=100 clinical letters from the Department of Medicine I, University Hospital Carl Gustav Carus Dresden. The clinical letters were selected from 100 random patients who have been treated between September 2004 and January 2023. A plethora of residents and senior physicians from internal medicine wrote the letters in different combinations. The letters’ length ranges from short reports to long letters and their medical focus lies on general internal medicine, gastroenterology, hematology and oncology.

They encapsulate free text, tables for laboratory results, endoscopic intervention reports, radiology reports and consultative reports from other specialties. All research procedures were conducted in accordance with the Declaration of Helsinki. Ethics approval was granted by the ethics committee of Technical University Dresden, reference number BO-EK-400092023.

#### Data preprocessing and Model selection

All 100 clinical letters were collected in PDF format. We extracted the text from the PDF and divided it into smaller text chunks of 4000 characters each. This chunk size was chosen due to the limited context window size for prompt and text chunk processing of the benchmarked LLMs and allowed processing with all models. For LLM inference, we set up llama.cpp, an opensource C++ library that allows inference of LLMs on local hardware.^18^ We tested a small (7 billion parameters (7B))^19^ and a larger model of Llama-2 (70 billion parameters (70B)),^20^ a llama-2 model that has previously been fine-tuned on German language, Llama-2 “Sauerkraut” 7B and 70B^21^ as well as Llama-3 with 8 billion parameter size (8B)^22^ and Llama-3 70B^23^. We extended the Llama models with Mistral 7B^24^ and Phi-mini.^25^ To enable use on comparatively low-resource hardware, we employed only quantized models, which are smaller than unquantized LLMs. The models were instructed to retrieve PII from each chunk of clinical letters programmatically. To ensure a consistent output structure, we used grammar-based sampling, an approach that allows predefining the output structure of the LLM in a json-formatted object via the llama.cpp framework. (Further details can be found in **Supplementary Materials**)

#### Definition and Extraction of Patient identifiers

In this paper, we defined PII as follows based on their relevance for clinical research and according to the findings in the clinical letters. PII include: patients first name, second name, birth name, date of birth, street, house number, zip code and patient id. This definition is not aligned with the Protected Health Information definition by the Privacy Rule of Health Insurance Portability and Accountability Act of 1996 (HIPAA) by the United States of America as this study was meant to demonstrate a proof of concept.

#### Evaluation of Model Results

To establish a reference ground truth for our experiments, all n=100 clinical letters were manually annotated by one medical doctor removing all personal identifying information (PII). Annotations were performed on the original PDF documents using the open source annotation tool “Inception” and exported in JSON-format.^26^ Annotated documents were exported and programmatically compared to the anonymized documents. Performance metrics for our results were calculated using macro averages, where metrics were computed individually for each clinical letter and subsequently averaged across all letters, yielding a comprehensive assessment of overall performance.

#### LLM-Anonymizer Pipeline Accessibility and Operation

We developed a fully automated pipeline, the LLM-Anonymizer, available on GitHub at https://github.com/KatherLab/LLMAnonymizer-Publication. The README file guides through setting up the pipeline, which can then be run from the terminal and accessed via browser. It simplifies data anonymization from real-world medical documents through a user-friendly interface with four main tabs (**Figure 1**):

1. **Preprocessing**: Upload raw data from various formats (e.g. PDF or TXT), preprocess it for use in the pipeline, and generate a zip file for storage.
2. LLM Information Extraction:

a. *Model Configuration:* Define LLM model parameters, select hyperparameters, adapt the prompt and specify items for identification in grammar.
b. *Run anonymization pipeline*: Hit “Run Pipeline” button. The model will be started and a progress bar tracks processing status and estimated time remaining.
3. **LLM Results**: Download processed files, including original, redacted, and pre-processed documents, stored in a zip file.
4. **Report Redaction**: Upload output zipped and annotated data sets for comprehensive reporting. Obtain global metrics, micro and macro scores, and a confusion matrix. Individual document analysis reveals false negative rates, ensuring thorough redaction of personally identifiable information. We included an option for fuzzy matching instead of exact matching, which allows flexibility in removing similar words that may account for spelling variations. More details on the pipeline steps can be found in the **Supplementary Materials**.

**Figure 1.**
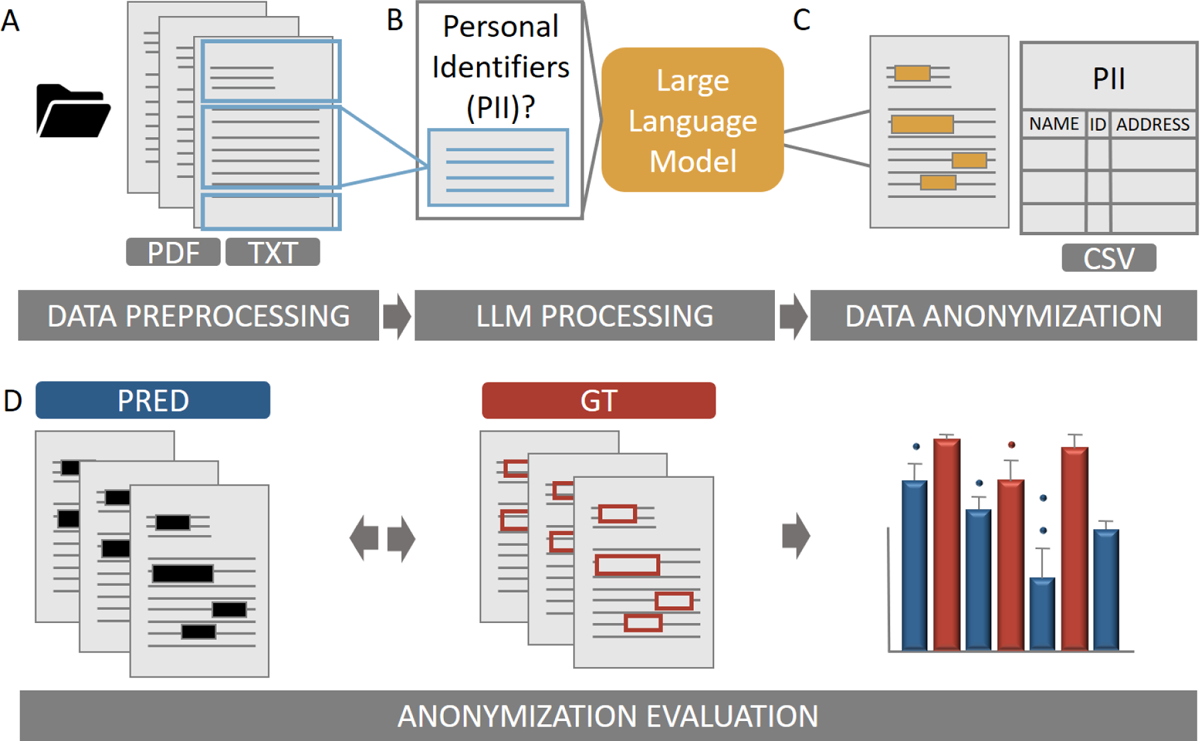
Anonymizer workflow. **A** Medical documents such as clinical letters in PDF and TXT format can be uploaded in the “data preprocessing” step. PDF input format is then converted into raw text, using optical character recognition (OCR) when necessary. The raw text files are then split into chunks of variable character size, which can be specified by the user. Different chunk sizes may be necessary due to the different context windows of the desired large language models (LLMs). **B** After preprocessing, the predefined LLM extracts personal identifiers from the document chunks and transfers them to a CSV file (LLM processing). **C** The personal identifiers are then hidden in a copy of the original input PDFs (Data anonymization). **D** For anonymization evaluation, each redacted document (PRED) can be compared to an annotated ground truth dataset (GT) and performance metrics (F1 score, accuracy, recall, precision, specificity, false positive rate and false negative rate) are calculated and displayed for each document as well as for all processed documents.

## Results

### LLMs can extract personal identifiers from clinical letters with high performance

In our pipeline, various locally deployable LLMs were prompted to extract the personal identifiers (patient name, first name, last name, birth name, birth date, address with street, street number and zip code, as well as patient id) from pre-processed clinical letters. The extracted information was redacted within the PDF using exact string matching. After this report redaction, we calculated performance metrics when comparing the pipeline results with our manual annotations (**Table 1**). We found high accuracy for all tested models (Llama 3 8B mean accuracy (mA) 97.3 ± 1.9%, Llama 3 70B mA 99.2 ± 0.9%, Llama 2 “Sauerkraut” 7B mA 99.10 ± 0.67%, Llama 2 “Sauerkraut” 70B mA 98.9 ± 0.91%, Mistral 7B mA 97.89 ± 2.30%). Highest recall (sensitivity) was achieved by Llama 3 models (**Figure 2 A and C**), with Llama 3 8B performing at the same level as Llama 3 70B, indicating a high fraction of PII being correctly redacted (sensitivity 8B 97.63 ± 4.28%, 70B 97.94 ± 6.11%). Mistral 7B and Llama 2 7B “Sauerkraut” showed the weakest sensitivities with 86.19 ± 13.05% and 76.61 ± 12.69% respectively. Llama 2 70B “Sauerkraut” also showed high performance with 90.37 ± 11.63%. The model with the highest precision was Llama 2 7B “Sauerkraut” with 76.68 ± 17.38%, followed by Llama 3 70B (74.34 ± 16.233%), Llama 2 70B “Sauerkraut” 68.53 ± 21.90%, Mistral 53.61 ± 22.36% and Llama 3 8B 43.79 ± 16.55% with the lowest precision.

**Figure 2.**
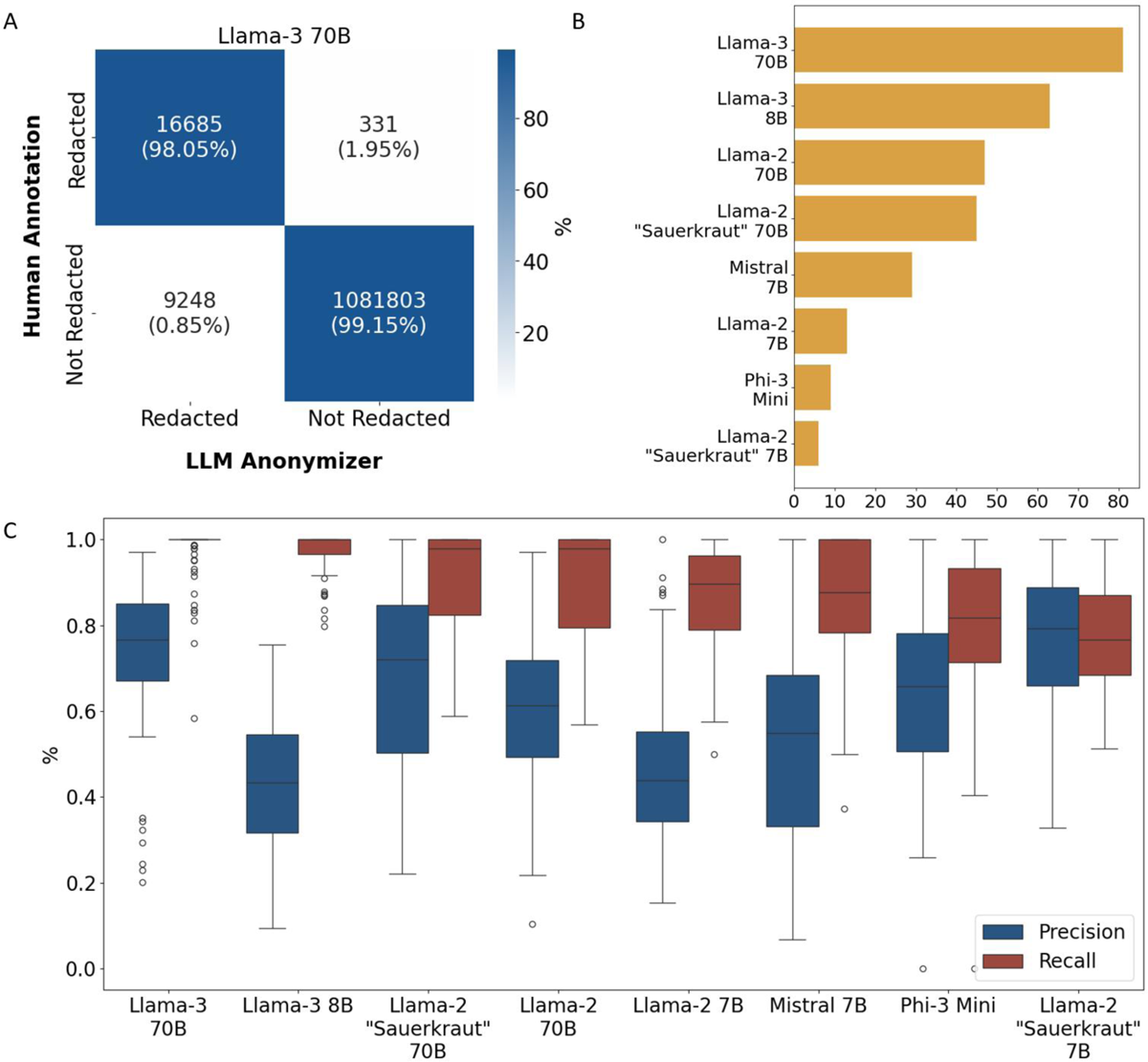
Model comparison for Anonymization of medical documents. **A** The confusion matrix presents the recall (sensitivity) and specificity of Llama-3 70B for de-identification of n=100 real-world clinical letters. The LLM-Anonymizer correctly redacted 98.05% of patient-sensitive identifiable information (character-wise), with only 1.95% of the information to be redacted being missed and 0.85% being redacted even if not necessary. **B** The Llama-3 models demonstrated the greatest effectiveness in the number of redacted letters, where no potential sensitive information was missing to be redacted. While Llama-3 70b did not miss any information in n=81 letters, the least effective model, Llama-2 “Sauerkraut” 7B, did not miss any information in less than 10 letters. **C** The box plots illustrate the distribution of the performance metrics precision and recall for all tested models. A high recall indicates the percentage of characters that have been correctly redacted, whereas the precision demonstrates the percentage of characters containing personal identifying information among all characters redacted.

**Table 1.** Performance Metrics of all tested models. Macro Scores of Mean, Standard Deviation (std) and Median are shown from the anonymization experiments with n=100 clinical letters compared to manually annotated data.

| model | metric | mean | std | median |
| --- | --- | --- | --- | --- |
| llama-2-70b-chat | specificity | 0.9873 | 0.0139 | 0.9906 |
|  | false positive rate | 0.0127 | 0.0139 | 0.0094 |
|  | false negative rate | 0.1021 | 0.1218 | 0.0208 |
|  | recall | 0.8979 | 0.1218 | 0.9792 |
|  | accuracy | 0.9858 | 0.0136 | 0.9887 |
| llama-2-7b-chat | specificity | 0.9802 | 0.0114 | 0.9821 |
|  | false positive rate | 0.0198 | 0.0114 | 0.0179 |
|  | false negative rate | 0.1319 | 0.1154 | 0.1040 |
|  | recall | 0.8681 | 0.1154 | 0.8960 |
|  | accuracy | 0.9782 | 0.0113 | 0.9800 |
| Meta-Llama-3-70B-Instruct | specificity | 0.9923 | 0.0093 | 0.9951 |
|  | false positive rate | 0.0077 | 0.0093 | 0.0049 |
|  | false negative rate | 0.0206 | 0.0611 | 0.0000 |
|  | recall | 0.9794 | 0.0611 | 1.0000 |
|  | accuracy | 0.9920 | 0.0092 | 0.9948 |
| Meta-Llama-3-8B-Instruct | specificity | 0.9737 | 0.0180 | 0.0180 |
|  | false positive rate | 0.0263 | 0.0225 | 0.0180 |
|  | false negative rate | 0.0237 | 0.0000 | 0.0428 |
|  | recall | 0.9763 | 1.0000 | 0.0428 |
|  | accuracy | 0.9736 | 0.9778 | 0.0178 |
| mistral-7b-instruct-v0.1 | specificity | 0.9810 | 0.0230 | 0.9863 |
|  | _false positive rate | 0.0190 | 0.0230 | 0.0137 |
|  | false negative rate | 0.1381 | 0.1305 | 0.1233 |
|  | recall | 0.8619 | 0.1305 | 0.8767 |
|  | accuracy | 0.9790 | 0.0227 | 0.9836 |
| Phi-3-mini-128k-instruct | specificity | 0.9912 | 0.0073 | 0.9935 |
|  | false positive rate | 0.0088 | 0.0073 | 0.0065 |
|  | false negative rate | 0.1879 | 0.1562 | 0.1824 |
|  | recall | 0.8121 | 0.1562 | 0.8176 |
|  | accuracy | 0.9880 | 0.0081 | 0.9900 |
| sauerkrautlm-70b-v1 | specificity | 0.9905 | 0.0089 | 0.9940 |
|  | false positive rate | 0.0095 | 0.0089 | 0.0060 |
|  | false negative rate | 0.0954 | 0.1155 | 0.0215 |
|  | recall | 0.9046 | 0.1155 | 0.9785 |
|  | accuracy | 0.9890 | 0.0091 | 0.9913 |
| sauerkrautlm-7b-v1 | specificity | 0.9948 | 0.0054 | 0.9965 |
|  | false positive rate | 0.0052 | 0.0054 | 0.0035 |
|  | false negative rate | 0.2339 | 0.1269 | 0.2335 |
|  | recall | 0.7661 | 0.1269 | 0.7665 |
|  | accuracy | 0.9907 | 0.0066 | 0.9928 |

### Llama 3 and large Llama 2 Models most effectively de-identify clinical letters

The false negative rate, which is defined as the proportion of personal identifiers where redaction was missed by the LLM-Anonymizer, was lowest for the Llama-3 70B-model, which achieved a FNR of 2.06% ± 6.11% (Macro Scores). This model also had the highest number of clinical letters without any personal identifiers that were missed during the redaction process (n= 81) (**Figure 2B**). The models Llama-3 8B (2.40% ± 4.32%) and Llama-2 70B “Sauerkraut” (9.63% ± 11.63%) exhibited the next best performance, with 63 and 45 letters, respectively, without any instances of missed redactions. Overall, the models in the Llama-3 group performed best at redacting sensitive information. This is illustrated in **Figure 3**, which shows the FNR of all labels per model. Larger models with more model parameters performed better with a FNR distribution towards 0. Llama-3 8B showed best performance among smaller parameter sized models.

**Figure 3.**
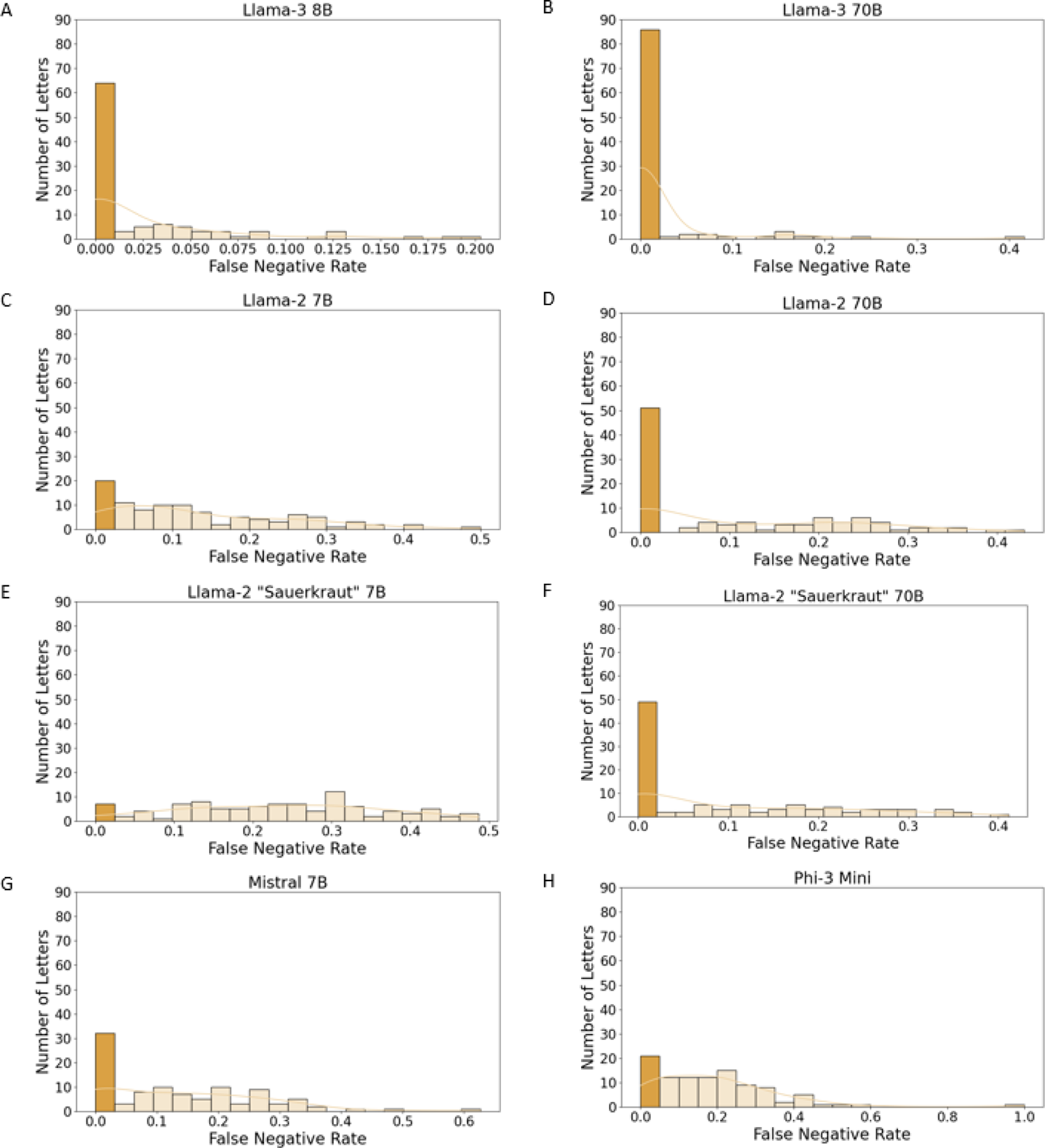
False negative rate per clinical letter. When anonymizing medical documents, the False Negative Rate (FNR) per doctor’s letter is critical to evaluate the effectiveness of de-identification. The y-axis represents the number of letters redacted by the anonymizer, while the x-axis represents the FNR. An FNR of 0 indicates that all human annotations were redacted in the clinical letters, which is highlighted in dark blue. Llama-3 models show the best FNR with a slight improvement from the smaller-(8 billion parameters, Panel **A**) to the larger model (70 billion parameters, Panel **B**). **C** and **D** show the results for Llama-2 models with 7 billion and 70 billion parameter size. **E** and **F** depict the FNR of llama-2 based models which were fine-tuned on German language “Sauerkraut”.^21^ **G** and **H** show the FNR of Mistral 7B and Microsoft’s Phi-3 model.

### Llama-3 outperforms at extracting personal identifiers across all categories

Patient first name and date of birth were consistently the most accurately extracted personal identifiers across all models. All information for the patient’s date of birth was de-identified with no missing characters in 99/100 letters for Llama-2 70B, Llama-2 7B, Llama-3 70B, and Llama-2 “Sauerkraut” 70B. Llama-3 8B and Mistral 7B identified it completely in 98/100 letters, Llama-2 “Sauerkraut” 7B in 97/100 and Phi-3 Mini in 87/100 letters. The label house number, on the other hand, proved to be more difficult for almost all models to identify correctly, with only 75/100 for Llama-3 70B, 19/100 for Llama-3 8B, 21/100 for Llama-2 70B, and below 20/100 for all other models. However, Llama-3 70B showed overall superior performance, achieving the lowest false negative rate across all identifier categories (**Figure 4**). While patient address and patient ID appeared most often only once in the dataset, patient name and date of birth appeared four to five times more frequently in the clinical letters (**Supplementary Figure 4**).

**Figure 4.**
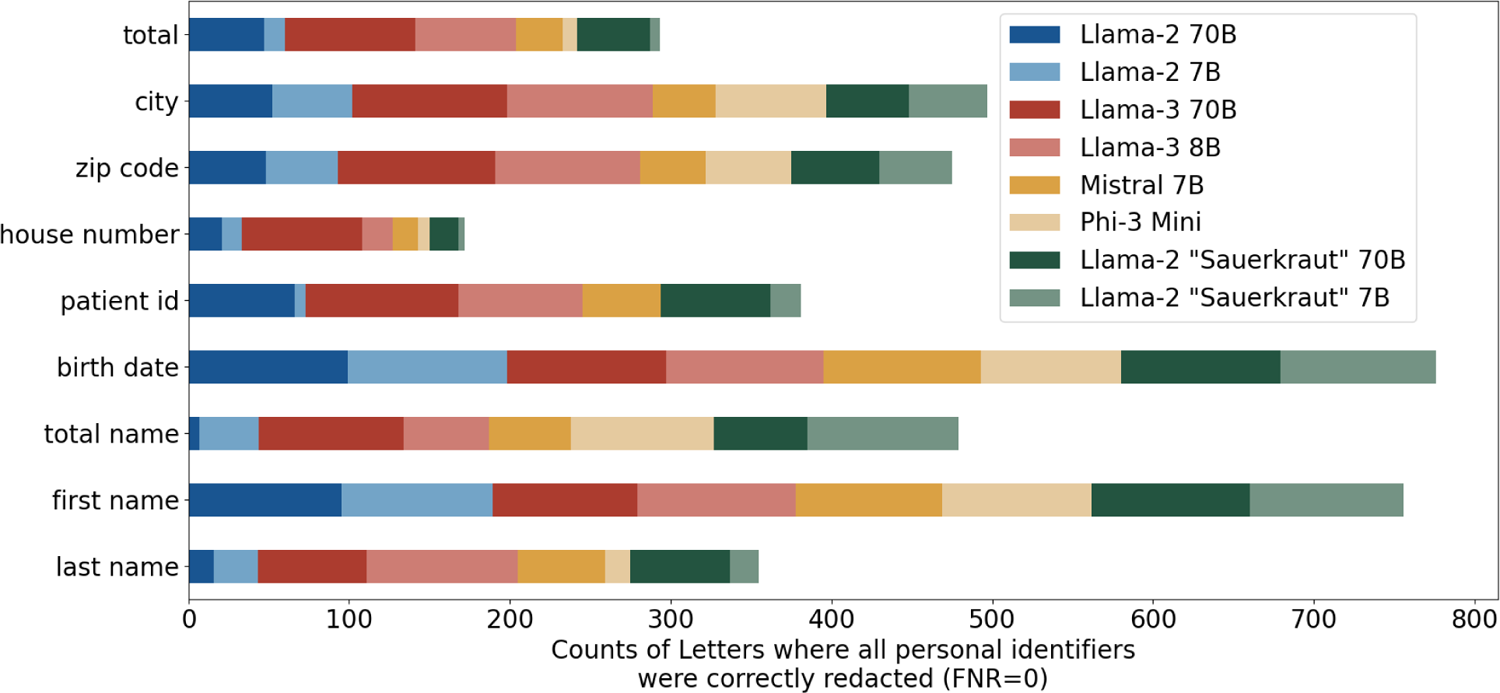
Zero False Negative Rates for All Models and Labels. The stacked bar chart shows the number of clinical letters with a false negative rate of zero for each individual label. First name and date of birth were extracted in nearly all letters for almost all models, while house number was more difficult to extract correctly for some models. The y-axis represents all labels for patient identifying information, such as first name, last name, date of birth, city, zip code, patient ID, birth name, and house number. The x-axis represents the number of letters for which all label occurrences were redacted (FNR=0). The bars are stacked to represent different models. Each color in the stacked bars corresponds to a specific model, including Llama-2 70B, Llama-2 7B, Llama-3 70B, Llama-3 8B, Mistral 7B, Phi-3 Mini, and Llama-2 “Sauerkraut” in the 70B and 7B variant.

### Llama-3 70B produces highly robust results

Since reproducibility is a relevant and critical issue for LLM based experiments, we repeated the experiments with the best model, Llama-3 70B, for three times. We found accuracy rates of 99.23, 99.13, 99.20 for all three model runs as well as similar false negative rates (1.65%, 0.91%, 2.06%) and recall (98.35%, 99.10%, 97.94%). The metrics across all clinical letters were tested for normality with the Shapiro-Wilk test and found to be not normally distributed. Therefore, we performed Kruskal-Wallis test to test for statistical significance between all three models’ results, which confirmed non-significance (accuracy comparison across models: pvalue=0.7039, recall comparison: p=0.5668, false-negative rates comparison: p=0.5668).

## Conclusion

Removal of PII from medical text data is critical to ensure the privacy of sensitive patient information, but it presents a formidable challenge to the scientific community for data use and sharing. Here, we benchmarked eight locally deployable LLMs for personal identifier extraction from 100 real-world clinical letters and their subsequent anonymization. Additionally, we present the LLM-Anonymizer, a comprehensive, automated pipeline that streamlines this process for secure anonymization of medical text data on local hardware and with a user-friendly interface, not requiring any programming knowledge.

Existing NLP NER approaches have demonstrated notable performance for anonymizing medical documents (rule based: recall 0.95, precision 0.93).^27^ Recent advancements in transformer models have showcased similar capabilities in various NLP tasks (precision and recall 0.94).^28^ LLMs like GPT-4 have proven to possess advanced anonymization skills.^29^, LLMs fine-tuned for medical purposes demonstrated a slightly worse performance (precision of 0.91, a recall of 0.95) in anonymizing medical documents.^27^ We are the first to show the high performance of inference from local LLMs’, specifically Llama-2 and −3 models, in extracting PII from medical documents, whereas others achieved only insufficient results: Liu et al. tried zero-shot medical text anonymization with GPT-4 and Llama models, but the Llama models failed to generate any relevant anonymization output for tested medical documents with an accuracy of 0.61. GPT-4 demonstrated an superior accuracy of 0.908 for implicit and 0.99 for explicit shot prompting on their synthetic medical dataset.^30^ Without exploiting the full potential of LLM capabilities, our LLM-Anonymizer achieved a comparably high and robust performance in anonymizing medical documents with zero shot inference for accuracy (Llama-3 70B 99.2) and recall (Llama-3 70B 97.9). In terms of accuracy, the LLM-Anonymizer performs equally to GPT-4, but the strength of our experiments is the local setup and the evaluation on highly various, real-world medical text data. These results demonstrate that the LLM-Anonymizer is not only competitive with all state-of-the-art (SOTA) NLP techniques in anonymizing medical documents, but also provides a locally deployable solution for real-world medical data.

The LLM-Anonymizer is designed with a flexible definition of de-identifying information, allowing a direct customization of entities within the prompt. This enhances its versatility, especially if specific personal identifiers are meant not to be redacted. It accepts different types of medical documents, for example PDF or TXT format, and can also process scanned documents using OCR. This takes into account the medical routine reality in which paper-based documents are often scanned for digitization. Additionally, it supports easy integration of new to come LLMs, facilitating the selection and exchange of the most suitable models without the need for specific training or fine-tuning, as it operates solely on LLM inference. Further optimization of performance is possible through in-context learning and further prompt engineering,^31^ which is explored by ongoing projects. Our LLM-Anonymizer enables the comparison of annotation quality against manually annotated datasets. It provides comprehensive metrics for the entire document set and individual documents. This allows research teams from different locations to upload their own ground truth to evaluate the LLM-Anonymizer on their specific dataset. The tool-assisted review of each document enables rapid assessment of redaction quality and the iteration of anonymization entities for the purpose of process improvement. This offers substantial flexibility and variability in addressing the critical task of anonymization of medical documents. We focused on recall and false negatives for feasibility assessment, because removing all personal identifiers completely is more critical than mistakenly removing non-personal information, as long as the original meaning of the text remains accessible. However, this approach may have resulted in a trade-off between precision and recall. Precision might have suffered because our redacted entity definitions were too fine-grained for some labels. For instance, the indiscriminate redaction of numerical entities, such as house numbers, when extracted independently rather than in context with other address details, could also mistakenly remove important lab values and other numerical data.

The degree of anonymization through redaction of PII is itself subject of research.^32^ Even if all common personal identifying attributes are omitted, the remaining information might still be sufficient to re-identify the individual.^33^ Additionally, the documentation of medical texts exhibits significant variability due to inconsistent record-keeping practices, variations in patient names and misspellings, which present challenges for traditional NER techniques in accurately identifying personal information.^34^ LLMs offer a special opportunity here: as they are capable of grasping concepts rather than just keywords, misspellings and inconsistent record-keeping practices do not pose a substantial concern. They may even be able to identify all possible identifying, more abstract entities in a multi-level approach, before these entities can be extracted for anonymization. This needs to be clarified in further research.

With regards to future improvements of our pipeline, next steps incorporate extending the evaluation towards all criteria as defined by the HIPAA, including personal information of treating physicians. Although our dataset is diverse and encompasses various medical document formats and content, its size remains constrained. The shape of medical documents including the revealed PII varies across hospitals and their informatics structure. Consequently, the generalizability of our findings needs to be evaluated on larger and more generalized, annotated datasets sourced from diverse documents of multiple healthcare facilities.

Despite numerous opportunities for improvement, we were able to show that LLMs are promising in identifying patient-sensitive data and present a valuable, easy-to-use, open-source tool that simplifies the anonymization of medical text data, facilitating its cross-site use.

## Code Availability Statement

We provide all code to use the LLM-Anonymizer and reproduce this study with own documents upon publication at: https://github.com/KatherLab/LLMAnonymizer-Publication

## Data Availability

The data used in this study contains patient sensitive information and will therefore not be published. We provide fictitious examples of clinical letters in the Supplement.

## Supplementary Materials

### Development of LLM-Anonymizer

The LLM-Anonymizer is a user-friendly, open-source software pipeline that allows LLM-based PII-extraction from documents and automatically retrieves this information from the documents for anonymization. Additionally, evaluation of the process performance can be executed. The pipeline is a Python Flask-based web app that builds on llama.cpp.^18^ Llama.cpp is an open-source C++ library that implements LLMs for efficiency and compatibility with a broader range of hardware, including personal computers with powerful GPUs. It allows LLM-inference locally without requiring access to proprietary systems or cloud resources. When llama.cpp is set up, the GitHub repository’s README file guides through all further steps. Anonymization Pipeline

1. Data preprocessing Export formats from clinical information systems vary widely among healthcare providers. The most common formats are documents in Portable Document Format (PDF) or standard document formats (e.g. DOC). The LLM anonymizer allows the upload of a variety of input formats and preprocesses the data for further processing in the anonymization pipeline. Supplementary Figure 1A shows the preprocessing interface. Longer documents may require partitioning into smaller chunks to accommodate the context window constraints of the target LLM. For this purpose, the desired chunk size should be specified before starting the preprocessing phase. Initiating the process is accomplished by clicking the “Preprocess Files” button. A progress bar indicates the progress and completion of the preprocessing procedure. Afterwards, a preprocessed zip file can be downloaded, which contains the original documents and the preprocessed raw text in a CSV file. Preprocessing is also able to read PDF scans or images, they are processed by OCR and stored separately.
2. LLM Information Extraction This tab serves the dual purpose of facilitating the extraction of personal identifying information (PII) and the automated anonymization of preprocessed medical documents through the use of an LLM (Supplementary Figure 1B). Initially, a preprocessed zip file must be uploaded, which enables flexible exchange of preprocessed files. Subsequently, the LLM must be configured. At this stage, a default prompt and grammar are provided, though they can be tailored to suit specific requirements. The model output must be in structured JSON format, which is achieved through the use of a grammar-based sampling approach. A default grammar for PII is predefined, however the user may specify other PII items in the grammar. The desired model can then be selected from a dropdown list, and the temperature parameter can be set (a low temperature, e.g., 0.1, is recommended for deterministic output). Clicking “Run Pipeline” uploads the model and initiates the anonymization process. It is possible to initiate multiple jobs in sequence, with each subsequent job queued and executed in succession. Upon completion of the process, the results may be obtained by clicking the “Download results” button.
3. LLM Results The additional tab “LLM Results” offers the possibility to download processed files separately. (Supplementary Figure 3)
4. Report Redaction The “Report Redaction” tab enables the evaluation of anonymization on a global scale (for the entire dataset) and on an individual level (for each document) if an annotated ground truth dataset is provided. The annotation dataset must be uploaded in the form of a zipped file containing UIMA CAS JSON 0.4.0-formatted annotation files. Following the upload of the output zipped and annotated data sets, the “Report Redaction” button should be executed (Supplementary Figure 2A). Once the progress bar indicates the completion of the report redaction process, the evaluation results can be reviewed by clicking the “View Report Redaction” button. This will display the global metrics, including the micro and macro scores, as well as a confusion matrix. The individual document analysis will reveal the false negative rates, enabling thorough redaction of each document where anonymization of PII was missed by the LLM-Anonymizer. Additionally, the option for fuzzy matching instead of exact matching has been included, allowing flexibility in removing similar words that may account for spelling variations. A datatable with all metrics (global and individual) can be downloaded as a CSV file.

### Model Benchmarking

#### Annotation

To establish a ground truth, n=100 clinical letters were annotated with the appropriate personal identifying information. Annotations were performed on the original PDF documents using the open source annotation tool “Inception”^26^. Annotated documents were exported in “UIMA CAS JSON 0.4.0”-file format. **Supplementary** Figure 4 shows counts per label for the manual annotations.

## Supplementary Figures

**Supplementary Figure 1.**
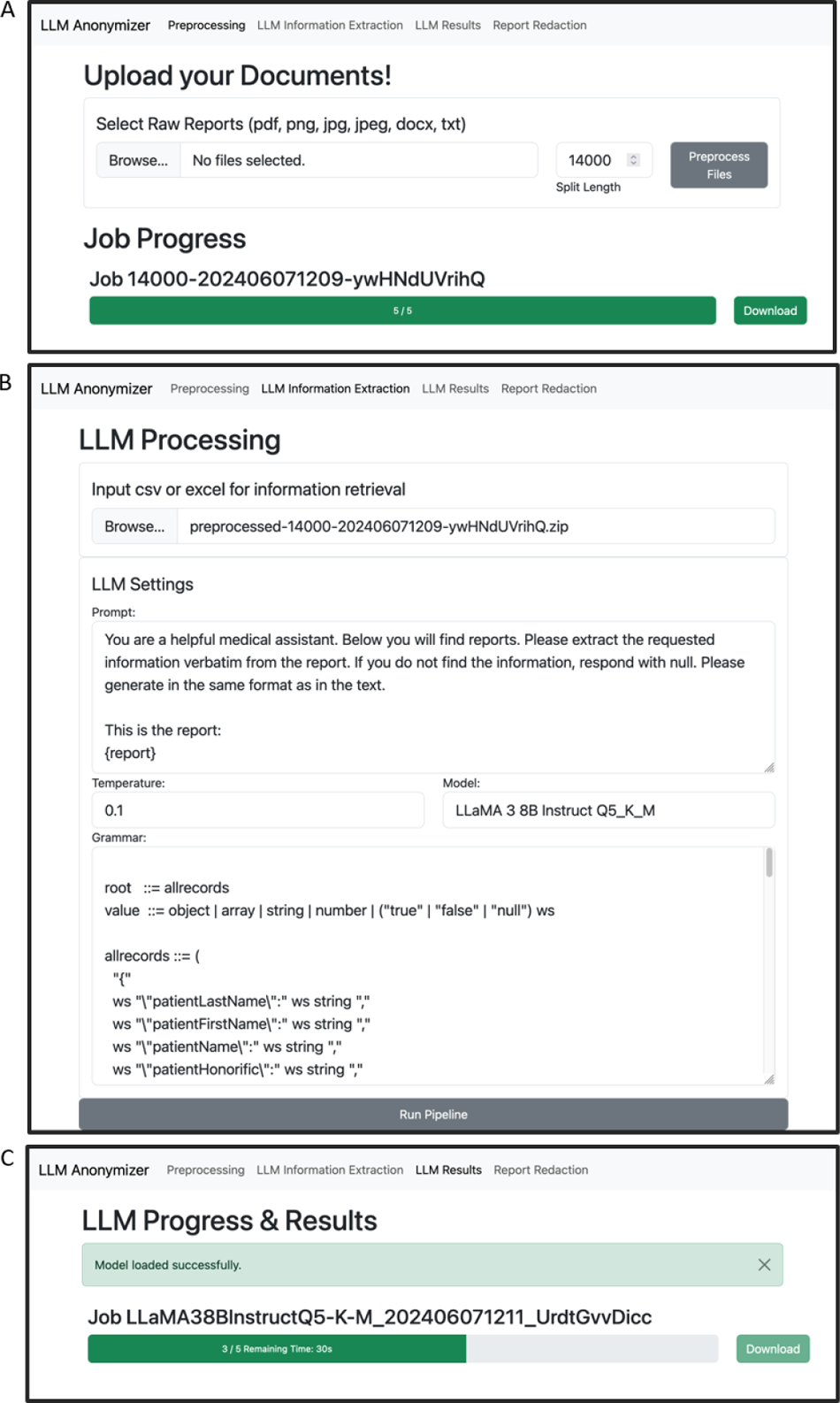
Anonymizer Preprocessing and Anonymization Graphical User Interface. The Anonymizer is a browser-based application that consists of four different tabs. A is the first tab, which allows data pre-processing. Raw reports can be uploaded in all common formats like PDF, DOCX, TXT. The documents are split into chunks of appropriate size and merged into a CSV. Clicking “Preprocess Files’’ will start the process and a progress bar will show the progress. Once the job is complete, it can be downloaded to the desired directory. B After preprocessing the data, the second tab “LLM Information Extraction’’ uploads a preprocessed CSV file. The Large Language Model, its settings, the prompt and the desired output format (grammar) must be specified. After clicking the “Run Pipeline’’ button, the process starts and a progress bar shows the progress.

**Supplementary Figure 2.**
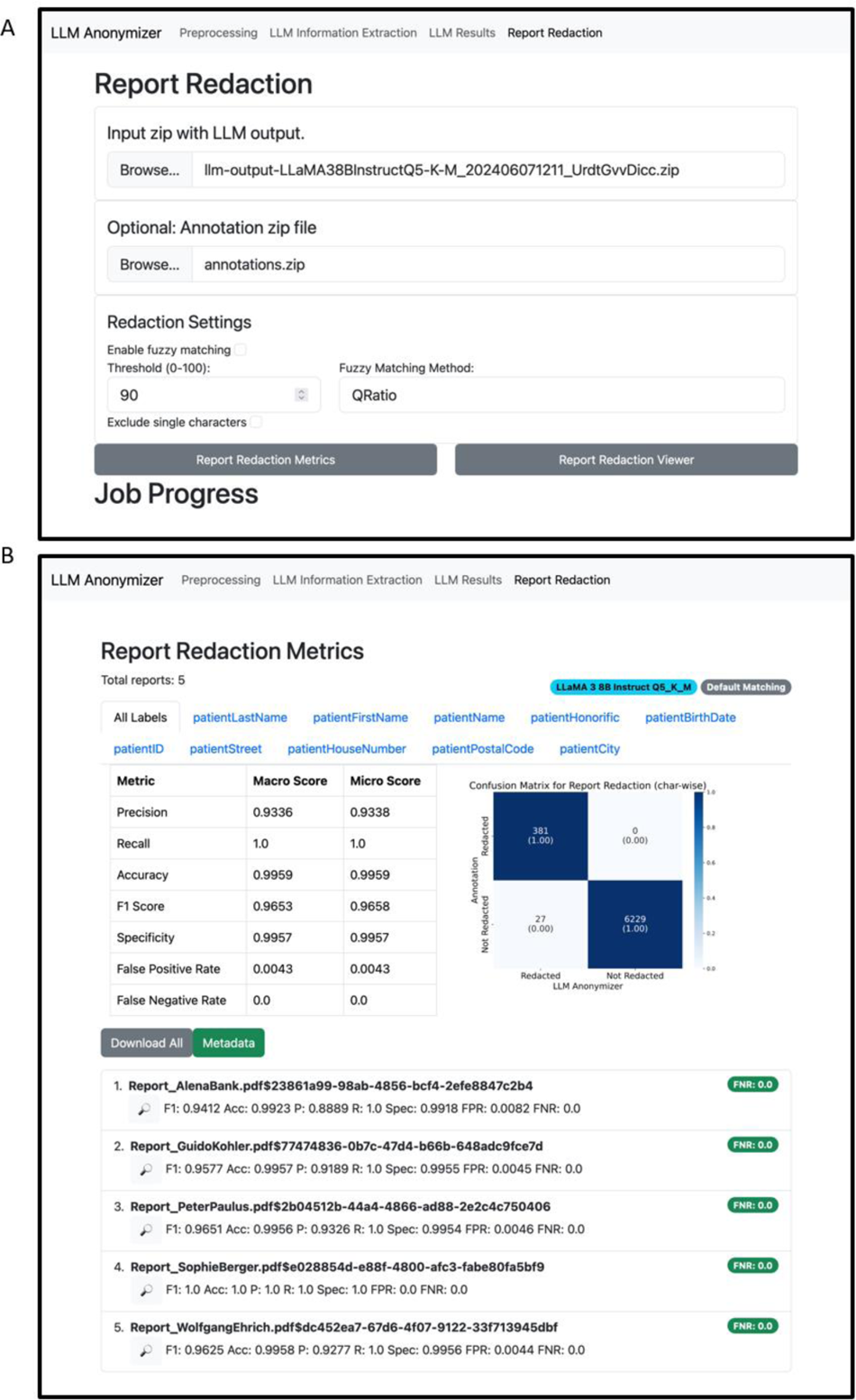
Anonymizer Report Redaction Graphical User Interface. A To view the redaction and redaction metrics of the medical documents, the output zip file must be uploaded. If an annotated dataset is available, it can also be uploaded. The LLM Anonymizer will mask the identified personal identifying information (PII) with keyword search. If desired, fuzzy matching can be enabled by clicking “Enable fuzzy matching”. All PII and similar words will be redacted, allowing to account for misspellings missed by the LLM. Clicking “Report Redaction Metrics” starts the process. B The output of this process is the Report Redaction Metrics overview, which displays global metrics for all uploaded documents with macro and micro scores. A Confusion Matrix shows the anonymization quality compared to the provided annotated dataset on a character-by-character basis. Additionally, all documents can be manually reviewed. The success rate of the anonymization process is indicated by the false negative rate, which describes the rate of characters missed by the anonymizer, for each document

**Supplementary Figure 3.**
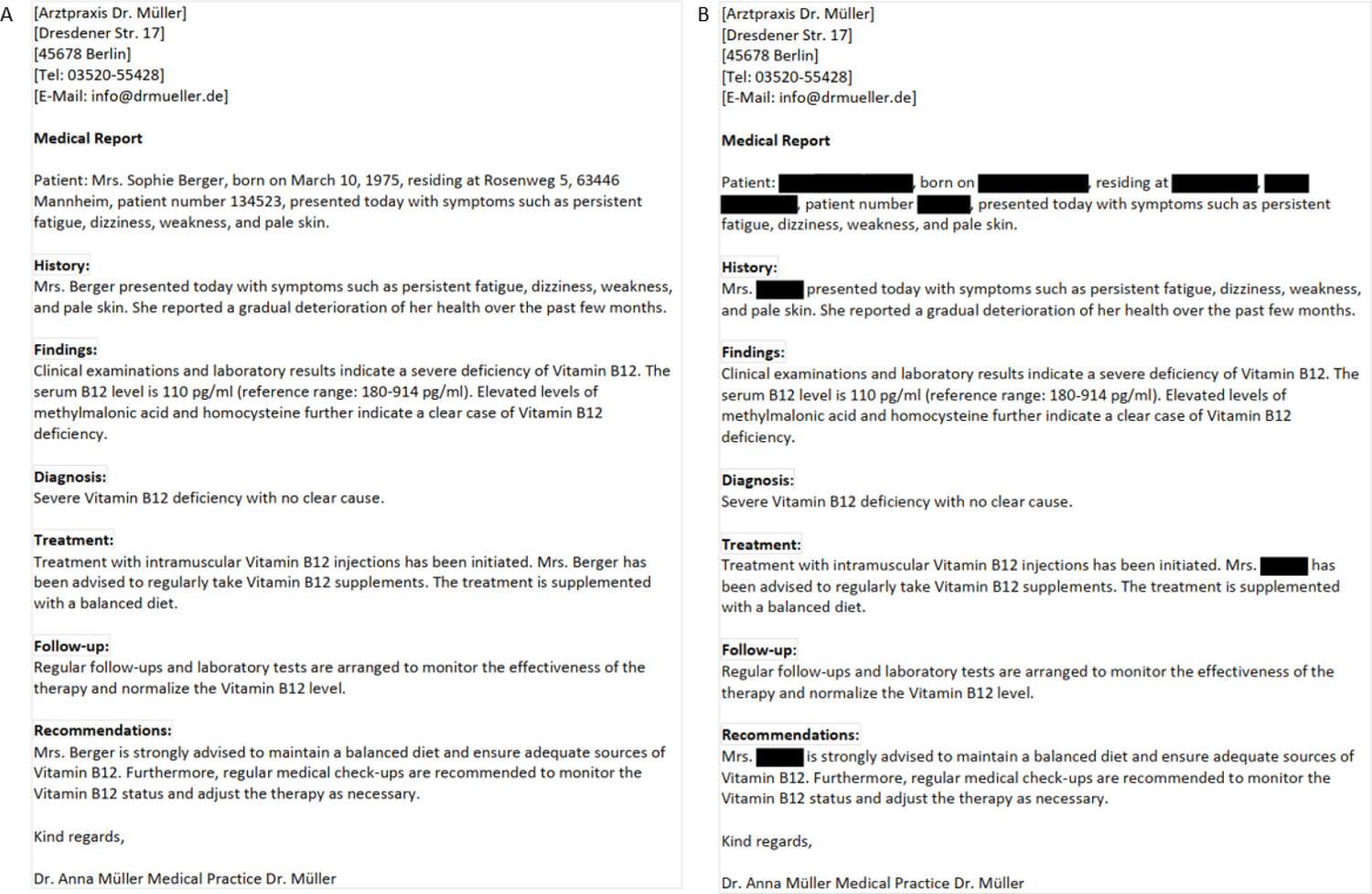
Clinical letter example. A Fictitious example of a clinical letter for display. Our dataset of real clinical letters was more complex, including longer letters and additional formatting elements, such as headers and footers with personal identifying information. The original data cannot be displayed for privacy reasons. B shows the document redacted by the LLM-Anonymizer.

**Supplementary Figure 4.**
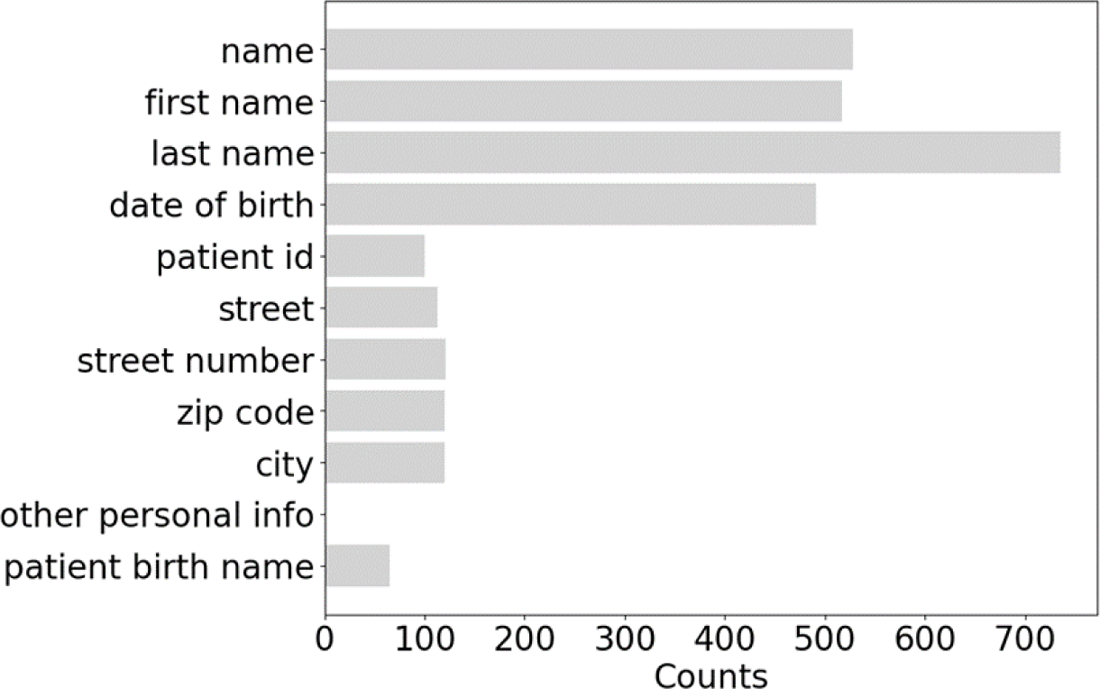
Counts of personal identifiers from ground truth annotations. Annotated labels are not evenly distributed across the dataset. The human-derived ground truth annotation revealed that patient name and date of birth are more frequently represented in our clinical letter dataset than patient ID, street, house number, zip code, city, and birth name. Other patient-identifying information that explicitly reveals personally identifiable information was only detected once by our ground truth annotator.

